# The Role of Decompressive Craniectomy following Microsurgical Repair of a Ruptured Aneurysm: Analysis of a South Australian Cerebrovascular Registry

**DOI:** 10.1101/2023.08.21.23294398

**Authors:** Tom J O’Donohoe, Christopher Ovenden, George Bouras, Seevakan Chidambaram, Stephanie Plummer, Andrew S Davidson, Timothy Kleinig, Amal Abou-Hamden

**Affiliations:** Department of Neurosurgery, Royal Adelaide Hospital, South Australia, Australia; University of Adelaide, South Australia, Australia; Department of Neurosurgery, Royal Melbourne Hospital, Victoria, Australia; Stroke Unit, Royal Adelaide Hospital, South Australia, Australia

## Abstract

**Background:** Decompressive craniectomy (DC) remains a controversial intervention for established or anticipated intracranial hypertension among patients with aneurysmal subarachnoid haemorrhage (aSAH).

**Methods:** We identified aSAH patients who underwent DC following microsurgical aneurysm repair from a prospectively maintained cerebrovascular registry and compared their outcomes with a propensity-matched cohort who did not. Logistic regression was used to identify predictors of undergoing decompressive surgery and post-operative outcome. The outcomes of interest were inpatient mortality, unfavourable outcome at first and final follow-up, NIS-Subarachnoid Hemorrhage Outcome Measure and modified Rankin Score (mRS) at first and final follow-up.

**Results:** A total of 246 consecutive patients with aSAH underwent microsurgical clipping of the culprit aneurysm between 01/09/2011 and 20/07/2020. Of these, 46 patients underwent DC and were included in the final analysis. Unsurprisingly, patients treated with DC had a greater chance of unfavourable outcome (p<0.001) and higher median mRS (p<0.001) compared with those who did not at final follow-up. Despite this, almost two-thirds (64.1%) of patients undergoing a DC had a favourable outcome at this time-point. When compared with a propensity-matched cohort who did not, patients treated with DC fared worse at all endpoints. Multivariable logistic regression revealed that the presence of intracerebral haemorrhage (ICH) and increased pre-operative mid-line shift were predictive of undergoing DC, and WFNS grade ≥ 4 and a delayed ischaemic neurological deficit requiring endovascular angioplasty were associated with an unfavourable outcome.

**Conclusions:** Our data suggest that DC can be performed with acceptable rates of morbidity and mortality, particularly among patients who present with lower grade aSAH. Further research is required to determine the superiority, or otherwise, of DC compared with structured medical management of intracranial hypertension in this context, and to identify predictors of requiring decompressive surgery and patient outcome.

## Introduction

Aneurysmal subarachnoid haemorrhage (aSAH) is frequently associated with raised intracranial pressure (ICP). ^1,2^ This can occur at the time of presentation, or in a delayed fashion, and has been associated with increased morbidity and mortality. ^2–7^ Therefore, although the management of raised ICP in patients with aSAH remains controversial, medical and surgical therapies are generally instituted with the intent of mitigating secondary brain injury. ^8,9^

Decompressive craniectomy (DC) is an effective intervention for the reduction of ICP and has been associated with improved glucose utilization and reduced cellular stress in patients with aSAH. ^10^ It has also been proposed to optimize the management of delayed cerebral ischaemia by permitting more vigorous hypertensive and hypervolaemic therapy, which, in the absence of decompression, may be associated with prohibitive elevations in ICP. ^4,11^ Despite this, previous experience with DC suggests unsatisfactory outcomes in patients with aSAH, with no significant difference observed in pooled rates of poor outcome between patients undergoing DC relative to matched controls at early or late follow-up in a recent meta-analysis. ^12^ However, although substantial heterogeneity precluded pooled subgroup analyses, it is likely that the outcome of patients undergoing DC varies significantly by the timing of, and indication for, decompression. ^12,13^ Both Buschmann et al ^3^ and Dorfer et al ^13^ have reported favourable modified Rankin Scale (mRS) scores in 59.4% and 34.8% of patients, respectively, who were undergoing DC for haematoma or oedema, as compared to 16.7% and 10.0% of patients having surgery for cerebral infarction secondary to delayed cerebral ischaemia. This distinction may also explain the observation that patients undergoing late DC are more likely to experience an unfavourable outcome when compared with those having earlier surgery. ^4,12,14^

Given that a number of authors have observed improved outcomes in patients undergoing earlier surgical decompression in patients with aSAH ^4,12,14^ and malignant cerebral infarction, ^15^ the identification of risk-factors associated with delayed intracranial hypertension may provide an opportunity for earlier selection of patients with aSAH who are likely to require DC.

This research sought to analyse a prospectively maintained Australian cerebrovascular registry of surgically treated ruptured aneurysms to; 1) describe the outcome of adult patients undergoing a DC for the presence, or anticipation of, intracranial hypertension following surgery, 2) compare outcomes between patients undergoing DC and a propensity matched cohort from the same registry who did not, 3) identify predictors of undergoing or requiring a DC and 4) identify predictors of poor outcome among patients undergoing decompressive surgery.

## Methods

### Study Design & Setting

This study comprised of a retrospective review of data from a prospectively maintained cerebrovascular database at The Royal Adelaide Hospital in Australia. It was approved by the Human Research Ethics Committee for the participating hospital (Ref: 14309).

### Participants

All adult (age ≥ 18) patients with aSAH who underwent microsurgical clipping with or without supratentorial DC were eligible for inclusion. Patients with significant coexisting traumatic brain injury, metastatic malignancy, pre-operative bilateral fixed and dilated pupils, intraoperative conditions that were deemed to be clearly non-survivable or those in whom the requirement for DC was not a direct consequence of the initial SAH, were excluded.

Patients undergoing DC were stratified into two subgroups; those who pre-emptively had their bone flap left out in the context of significant intracerebral haemorrhage (ICH) or the presence or anticipation of post-operative cerebral oedema following microsurgical aneurysm treatment or those who underwent rescue DC, either as a primary procedure, or as an extension of a previous craniectomy, for refractory intracranial hypertension following microsurgical aneurysm repair.

A composite of those patients who underwent pre-emptive DC and required post-operative neuromuscular blockade, hyperosmolar therapy or barbiturate coma for management of ongoing intracranial hypertension, and patients who underwent rescue DC, were compared with patients undergoing microsurgical clipping for ruptured cerebral aneurysms without primary or delayed DC.

### Data Extraction

Routine demographic and baseline clinical data were collected, including; age, gender, WFNS grade, aneurysm location, presence and volume of ICH at presentation, aneurysm re-rupture prior to treatment, delayed cerebral ischaemia requiring endovascular management, presence and extent of pre-operative midline shift (MLS), presence of unilateral fixed and dilated pupil, and mean ICP in 24 hours prior to decompression.

Operative information, including craniectomy type (hemicraniectomy or bifrontal craniectomy), size, and the time and indication for decompression were also obtained. Intracerebral haematoma volumes and craniectomy size were estimated from 5mm serial axial computed tomography voxels of the brain. Haematoma volume was calculated using the previously described ABC/2 method ^16^ and craniectomy size was estimated by the previously published equation of Tanrikulu ^17^ and Schur ^18^.

Outcome measures include overall and inpatient mortality, mRS at six- and twelve-months, and at last follow-up, tracheostomy, gastrostomy, overall length of hospital stay, discharge disposition and NIS-subarachnoid haemorrhage outcome measure (NIS-SOM). Discharge to institutional care included any discharge to a nursing facility, extended care facility, or hospice, but not transfer to rehabilitation or another acute care facility. Analysis of tracheostomy, gastrostomy, length of stay and discharge disposition were performed for surviving patients, in order to differentiate these endpoints from mortality. The NIS-SOM defines a poor outcome as in-hospital mortality, tracheostomy and gastrostomy placement, or discharge to institutional care and has been demonstrated to have 80% and 91% agreement with an mRS score of >2 and >3, respectively. ^19^ A mRS of 4-6 was classified as an unfavourable outcome. Primary DC was defined as any surgical decompression that occurred within 24-hours of admission, while secondary DC included all patients who underwent surgery after this time point.

### Statistical Analysis

Descriptive categorical data were summarised using absolute values and proportions, with numerical data being expressed according to their distribution. Dichotomous variables were assessed for independence using either a Pearson Chi^2^ test or Fisher’s Exact test depending on the expected cell frequencies.

Comparisons of central tendency between sets of continuous data were based on distribution. Normally distributed data were compared between groups with an independent two-sample t-test, whereas a Mann-Whitney test was used to compare non-parametric data, ordinal data or parametric data with significant outliers. Mean values are presented with standard deviation, while median values are accompanied by the 25^th^ and 75^th^ quartiles. For the purpose of ordinal analyses, mRS scores of 5 and 6 were combined.

Models were created to assess for predictors of the following outcome measures: dichotomised mRS at first follow up; ordinal mRS at first follow up; NIS-SOM; inpatient mortality; dichotomised mRS at final follow up; and ordinal mRS at final follow up. Binomial logistic regression was used for dichotomous measures, whereas ordinal logistic regression was utilised for ordinal or continuous variables. The following variables were assessed for a relationship with each outcome measure using a best subsets regression procedure: requirement of DC (yes/no), age (categorised into < 65 and ≥ 65), WFNS grade, dichotomised WFNS grade (WFNS <4 and WFNS ≥ 4), aneurysm location, presence of ICH, presence of a delayed ischaemic neurological deficit requiring endovascular management, extent of pre-operative MLS, presence of a unilaterally fixed and dilated pupil, aneurysm and craniectomy size. The model that minimised Aikaike’s Information Criterion ^20^ was selected.

A pseudo-R^2^ was calculated for each model using McFadden’s R^2^ method. Once each model was chosen, the dataset was split into a training and test set, and cross-validation resampling was utilised to assess its performance. Model specificity, sensitivity, receiver-operator characteristic (ROC) curves and the area under the ROC curve were created by averaging sensitivities and specificities across all folds. Overall model odds radios and confidence intervals were calculated by fitting the chosen model to the entire dataset.

Propensity matching was conducted to identify patients with a similar age, gender and presenting WFNS grade among those who did and did not undergo DC. A 1:1 matching protocol without replacement (‘nearest neighbour’ method) was utilised to generate the matched cohort.

Statistical analyses were conducted with Statistical Package for the Social Sciences (SPSS) Version 29 (IBM Corporation, Armonk, NY, USA) and R v 4.2.0 (R Foundation for Statistical Computing, Vienna, Austria). Graphs were produced using GraphPad Prism 9.0 (GraphPad Software Inc. Ca, USA) and Microsoft Excel 2016 (Microsoft Corporation, WA, USA). A p-value of <0.05 was considered to be statistically significant.

## Results

### Participants

A total of 246 consecutive patients with aSAH underwent microsurgical clipping of the culprit aneurysm between 01/09/2011 and 20/07/2020. Of these, 49 patients were treated with DC (46 primary and 3 secondary) at a mean of 8.9 (±27.5) hours after admission. Three patients who underwent DC were excluded from the analysis; two who were deemed to have clearly non-survivable pathology at the time of surgery and one who underwent DC due to a post-operative haematoma rather than as a consequence of the index haemorrhage. The vast majority (93.5%) of patients underwent a pre-emptive DC, while two patients (4.4%) undergoing delayed DC did so due to residual IPH/SDH with mass effect and one (2.2%) received surgery to alleviate mass effect secondary to a periprocedural infarction. Ten patients (21.7%) who received DC required post-operative medical management of raised ICP.

### Baseline Characteristics

The mean age (± SD) of participants undergoing DC was 51.0 years (±11.6) and 56.5% were female. The median (IQR) WFNS grade was 4 (2.0 – 5.0) and approximately one-quarter (26.1%) of patients had a fixed and dilated pupil prior to surgery. A majority (80.4%) of patients had an associated intracerebral haemorrhage with a median volume of 22.2cm^3^ (12.4 – 32.7). The median pre-operative MLS was 5.8mm (1.3 – 8.0) and over half (58.7%) of patients presented with a WFNS grade 4 or 5 SAH.

Given that this is a surgical series, all aneurysms were located on the middle cerebral (MCA), anterior communicating (ACOM), posterior communicating (PCOM) or anterior cerebral (ACA) arteries, and had a mean diameter of 8.3mm (± 3.9). The mean DC size was 57.8cm^2^ (±21.5) and ten patients (21.7%) developed a delayed ischaemic neurological deficit that required endovascular management. The baseline clinical and demographic characteristics of included participants are summarised in Table 1.

**Table 1:** Baseline Clinical and Demographic Characteristics of Included Participants Undergoing Decompressive Craniectomy.

| Baseline Demographic Data |  |  |  |
| --- | --- | --- | --- |
| Age [$\bar{x}$, {$\sigma$}] | | 51.0 (±11.6) | |
| Male:Female [n (%)] |  | 20:26 (43.5:56.5) |  |
| Baseline Clinical Data |  |  |  |
| WFNS Grade [n (%)] | | Aneurysm Size (mm) [$\bar{x}$, {$\sigma$}] | 8.3 (±3.9) |
| WFNS 1 | 8 (17.4) | Aneurysm Location [n (%)] |  |
| WFNS 2 | 7 (15.2) | MCA | 35 (76.1) |
| WFNS 3 | 4 (8.7) | ACOM | 3 (6.5) |
| WFNS 4 | 11 (23.9) | PCOM | 6 (13.0) |
| WFNS 5 | 16 (34.8) | ACA | 2 (4.4) |
| mFisher Grade [n (%)] | | ICH Volume [$M$ (IQR)] | 22.2 (12.4–32.7) |
| mFisher 1 | 2 (4.4) | Primary: Secondary DC [n (%)] | 43 (93.5):3 (6.5) |
| mFisher 2 | 4 (8.7) | DC Size ([$\bar{x}$, {$\sigma$}] (cm<sup>2</sup>)) | 57.8 (±21.5) |
| mFisher 3 | 9 (19.6) | Time of DC [$\bar{x}$, {$\sigma$}] (hours) | 8.9 (±27.5) |
| mFisher 4 | 31 (67.4) | Indication for DC [n (%)] |  |
| Presence of Unilateral Mydriasis [n (%)] | 12 (26.1) | Pre-emptive | 43 (93.5) |
| Extent of Pre-operative MLS (mm) [$M$ (IQR)] | 5.8 (1.3–8.0) | Residual IPH/SDH with oedema | 2 (4.4) |
| Vasospasm Requiring EV Mx [n (%)] | 10 (21.7) | Procedure related infarction | 1 (2.2) |
| ICH at Presentation [n (%)] | 37 (80.4) | Requirement for Post-DC ICP Mx [n (%)] | 10 (21.7) |

### Overall Outcome

The median time of first follow-up was significantly later among those who underwent a DC compared with those who did not (122.5 days vs 70 days, p<0.001). Unsurprisingly, the proportion of patients with a favourable outcome at this time-point was significantly lower in those undergoing a DC (57.8%) compared with those who did not (93.0%) (p<0.001) and a greater proportion of DC patients had a positive NIS-SOM result (41.3% vs 11.0%, p<0.001). Patients undergoing DC also had a higher mRS at first follow-up compared with those who did not (p<0.001), and a higher proportion of these patients died during their admission (p=0.04). The median time of final follow-up was similar between groups (p=0.06).

There was persistence of previously observed differences in functional outcome at final follow-up, with those undergoing a DC having a lower likelihood of favourable outcome (p<0.001) and a higher median mRS (p<0.001) compared with those who did not (Table 2 and Figure 1). Despite this, DC was not universally associated with poor outcome. Indeed, almost two-thirds (64.1%) of patients undergoing a DC had a favourable outcome at final follow-up. Reasonable rates of favourable outcome were also observed among patients with poor clinical grade (WFNS ≥ 4) who underwent DC, with 12 (44.4%) experiencing a favourable outcome at delayed follow-up.

**Figure 1:**
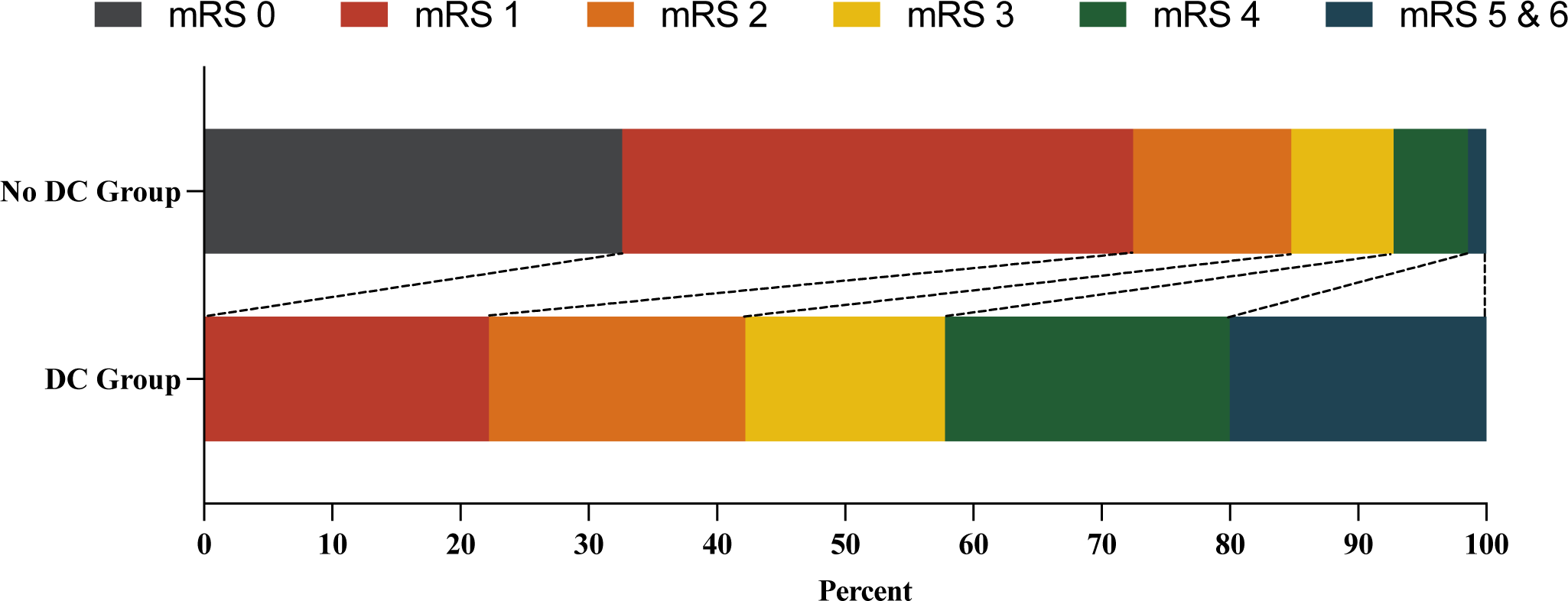
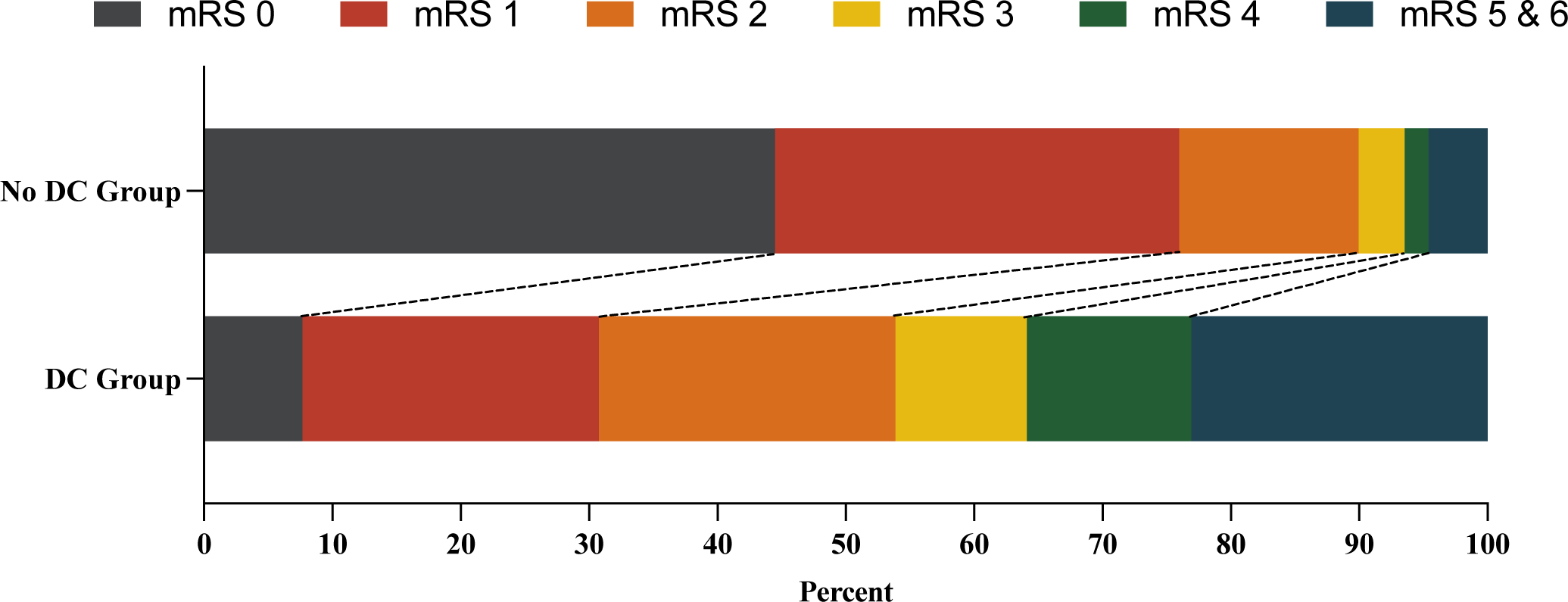
a: Stacked Bar Chart of Modified Rankin Scale (mRS) Scores at 1^st^ Follow-up Comparing Those who did and did not Undergo Decompressive Craniectomy b: Stacked Bar Chart of Modified Rankin Scale (mRS) Scores at Final Follow-up Comparing Those who did and did not Undergo Decompressive Craniectomy

**Table 2:**
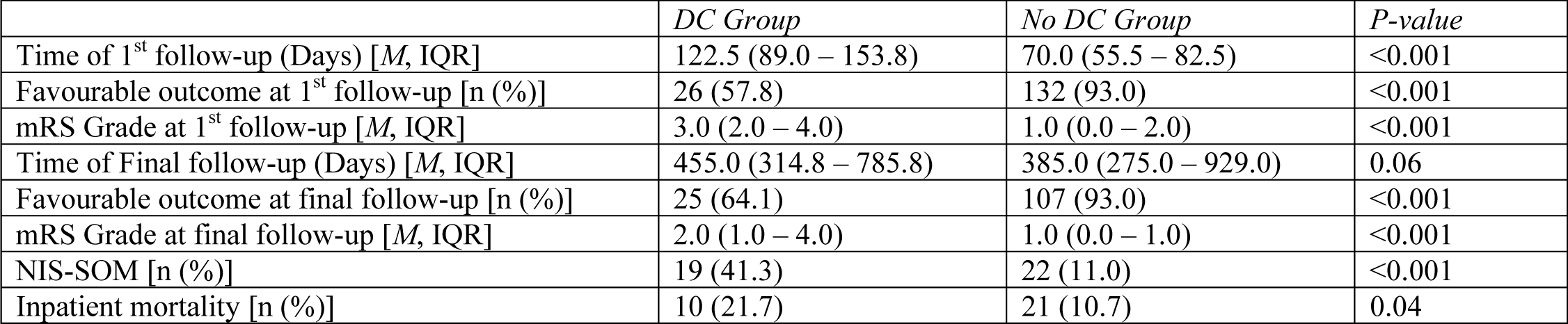
Comparison of Outcome Between Those Who Underwent Decompressive Craniectomy and Those Who Did Not.

### Propensity Matching

Outcomes were compared between patients undergoing DC and a propensity matched cohort who did not. The variables included in the model were gender, age and presenting WFNS grade. Patients were well matched for age and gender. Those in the matched DC cohort presented with a higher median WFNS grade (4.0 (2.0 – 5.0)) than matched controls (3.0 (2.0 – 4.0)), although this was not statistically significant (p=0.06). Patients treated with DC fared worse than their matched counterparts at all endpoints, with significantly higher rates of unfavourable outcome of first and final follow-up, inpatient mortality and NIS-SOM positive outcome (Table 3 & Figure 2).

**Table 3:** Comparison of Baseline Characteristics and Outcome Between Those Who Underwent Decompressive Craniectomy and Propensity-Matched Cohort Who Did Not.

|  | *DC Group* | *Matched Control* | *P-value* |
| --- | --- | --- | --- |
| **Baseline Characteristics** |  |  |  |
| Age [$\bar{x}$, {$\sigma$}] | 51.0 (11.7) | 52.9 (14.5) | 0.5 |
| M:F [n (%)] | 20 (44.4):25 (55.6) | 24 (53.3):21 (46.7) | 0.55 |
| WFNS Grade [*M*, IQR] | 4 (2.00 – 5.00) | 2 (2.0 – 4.0) | 0.06 |
| Vasospasm Requiring Endovascular Mx [n (%)] | 10 (22.2):35 (77.8) | 9 (20.0):36 (80.0) | 1 |
| **Outcome** |  |  |  |
| Unfavourable outcome at 1<sup>st</sup> follow-up [n (%)] | 19 (45.2) | 6 (14.3) | 0.004 |
| Unfavourable outcome at final follow-up [n (%)] | 14 (38.9) | 2 (6.3) | 0.002 |
| Inpatient mortality [n (%)] | 9 (21.4) | 0 (0.0) | 0.002 |
| NIS-SOM [n (%)] | 17 (40.5) | 6 (14.3) | 0.01 |
| mRS Grade at 1<sup>st</sup> follow-up [*M*, IQR] | 3.0 (2.0 – 4.0) | 1.0 (1.0 - 2.0) | <0.001 |
| mRS Grade at final follow-up [*M*, IQR] | 2.5 (1.8 – 4.5) | 1.0 (0.0 – 1.3) | <0.001 |

**Figure 2:**
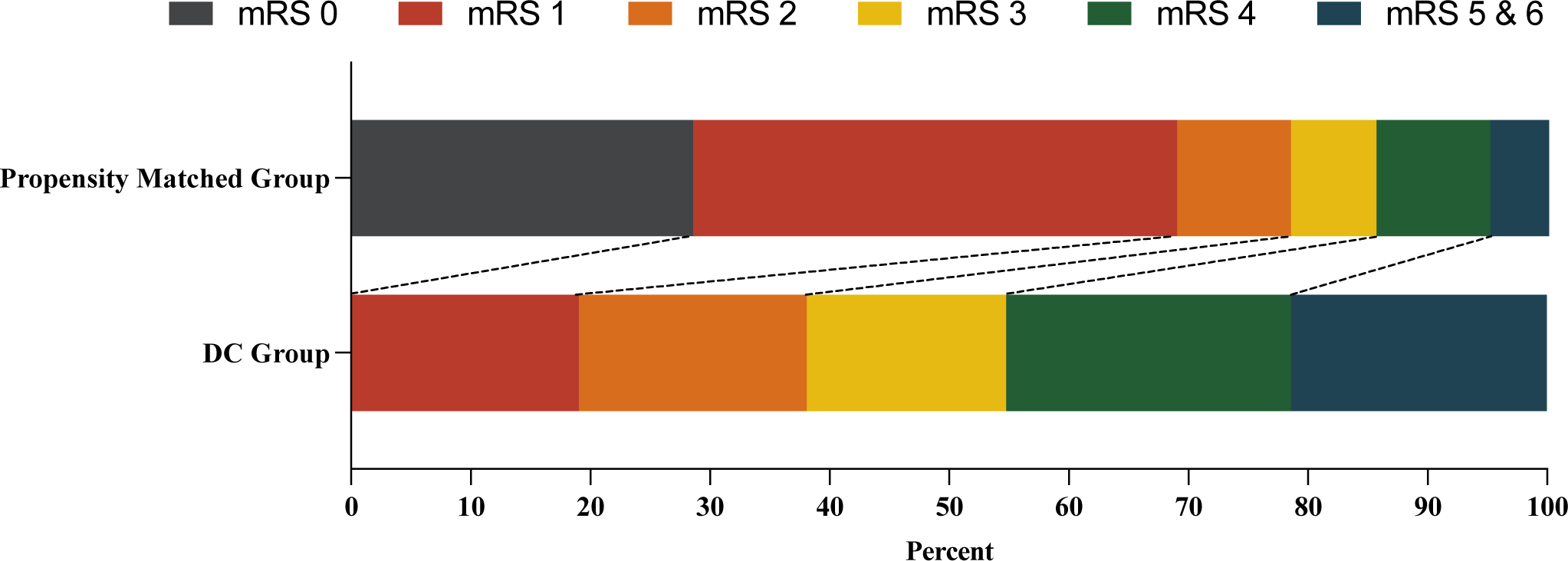
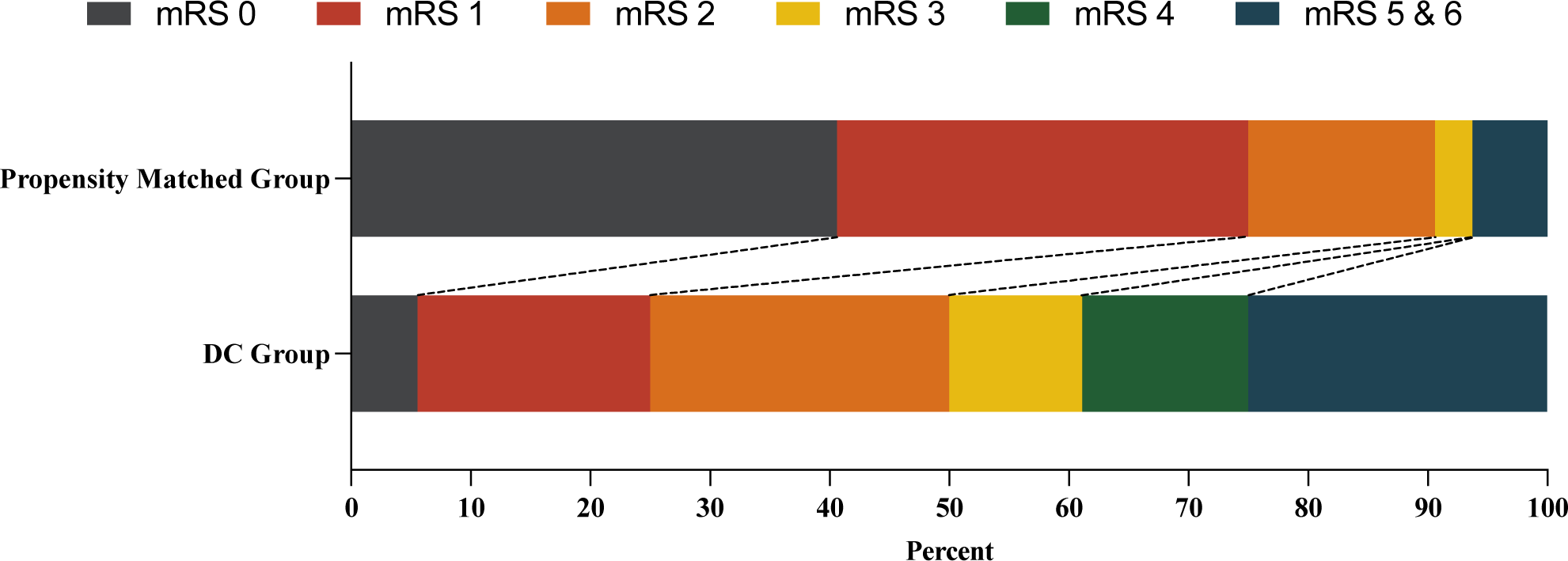
a: Stacked Bar Chart of Modified Rankin Scale (mRS) Scores at 1^st^ Follow-up Comparing Those who Underwent Decompressive Craniectomy with a Propensity Matched Cohort who did not b: Stacked Bar Chart of Modified Rankin Scale (mRS) Scores at Final Follow-up Comparing Those who Underwent Decompressive Craniectomy with a Propensity Matched Cohort who did not

### Predictors of Undergoing and Requiring DC

Given the retrospective nature of this research and anticipated variability in indications for DC between surgeons, we sought to identify a subset of those who underwent decompressive surgery, and had ongoing intracranial hypertension, as having unequivocally ‘required’ the DC. This subgroup was defined as a composite of patients who underwent a primary DC and required post-operative neuromuscular blockade, hyperosmolar therapy or barbiturate coma for management of ongoing intracranial hypertension, and patients undergoing secondary DC. Multivariable logistic regression was conducted to determine factors that were predictive of undergoing and requiring a DC. This analysis revealed that the presence of ICH (OR 14.7, 95% CI 4.4 – 48.8, p<0.001) and MLS >5mm (OR 5.1, 95% CI 1.5 – 17.5, p=0.01) were both predictive of undergoing DC (Figure 3). Only 13 patients reached criteria for requiring a DC and, given the small sample size, there were no predictive factors identified for this outcome.

**Figure 3:**
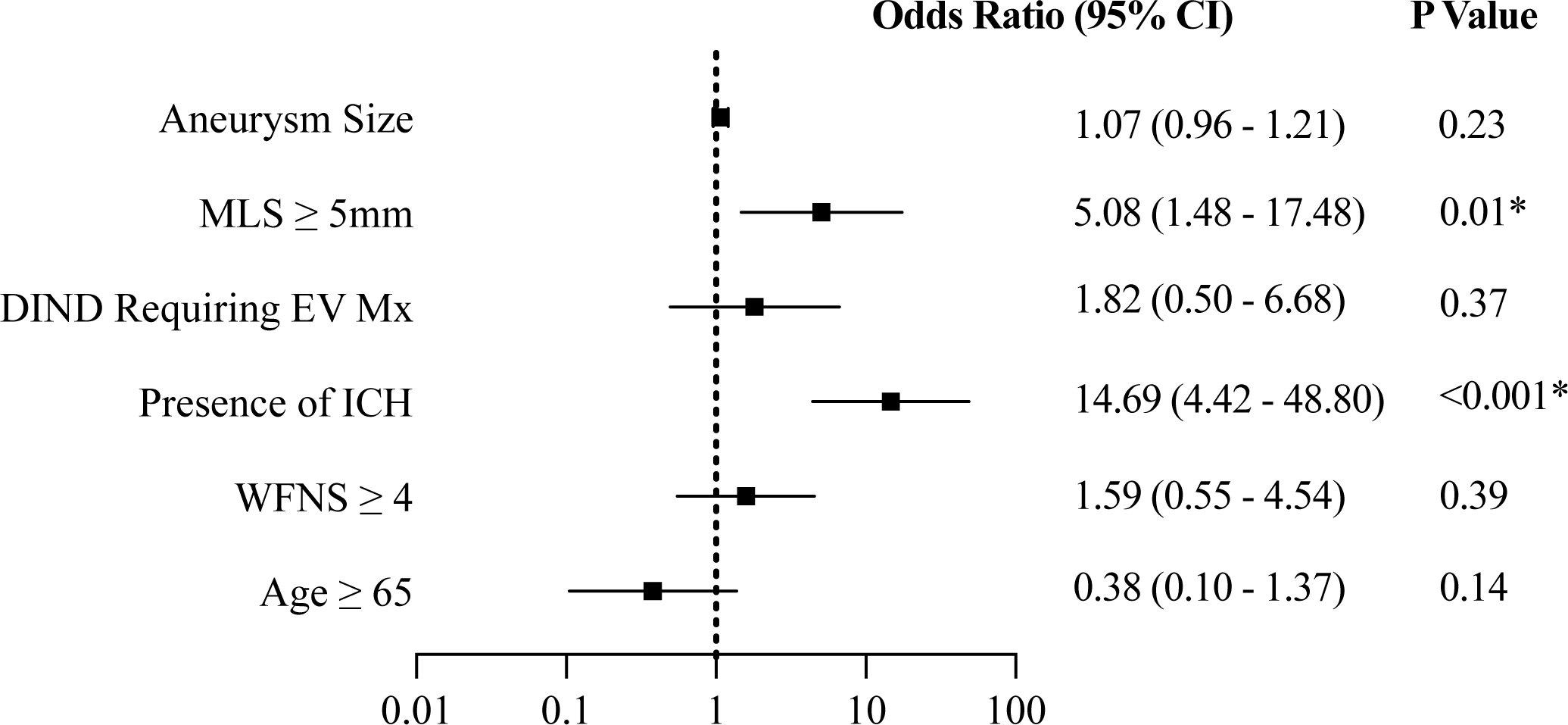
Independent Predictors of Undergoing Decompressive Hemicraniectomy. MLS: Midline Shift; DIND: Delayed Ischaemic Neurological Deficit; EV: Endovascular; Mx: Management; ICH: Intracerebral Haemorrhage; WFNS: World Federation of Neurosurgical Societies

### Outcome Prediction

Multivariable logistic regression demonstrated that WFNS grade ≥ 4 was associated with unfavourable outcome at first follow-up (Figure 3) and that patients requiring endovascular angioplasty for management of a delayed ischaemic neurological deficit were more likely to have an unfavourable outcome at first follow-up and a positive NIS-SOM result (Figures 4 & 5). No association was observed between the presence of pre-operative MLS ≥ 5mm, unilateral fixed and dilated pupil, aneurysm size or craniectomy size and either endpoint.

**Figure 4:**
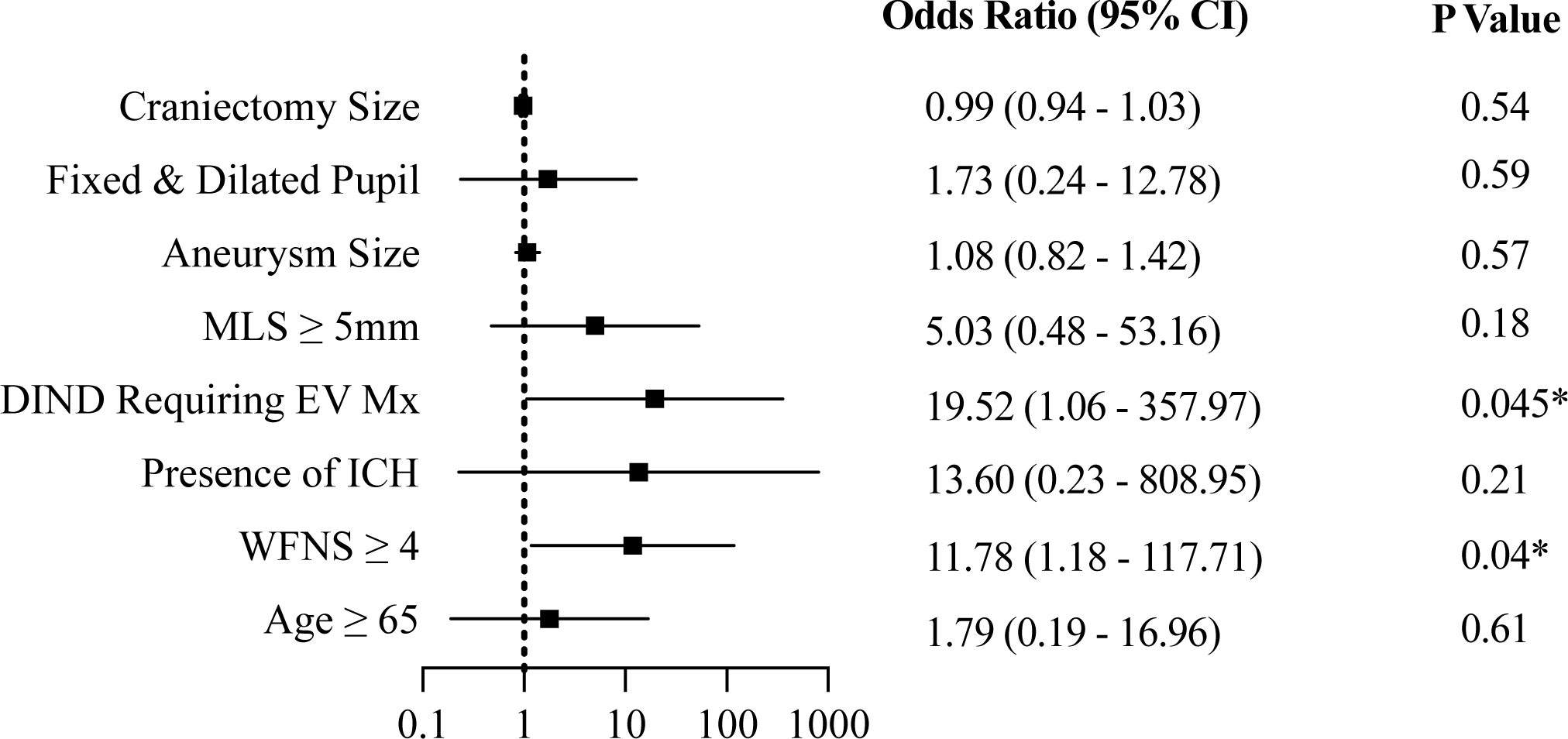
Independent Predictors of Unfavourable Outcome at 1^st^ Follow-up. MLS: Midline Shift; DIND: Delayed Ischaemic Neurological Deficit; EV: Endovascular; Mx: Management; ICH: Intracerebral Haemorrhage; WFNS: World Federation of Neurosurgical Societies

**Figure 5:**
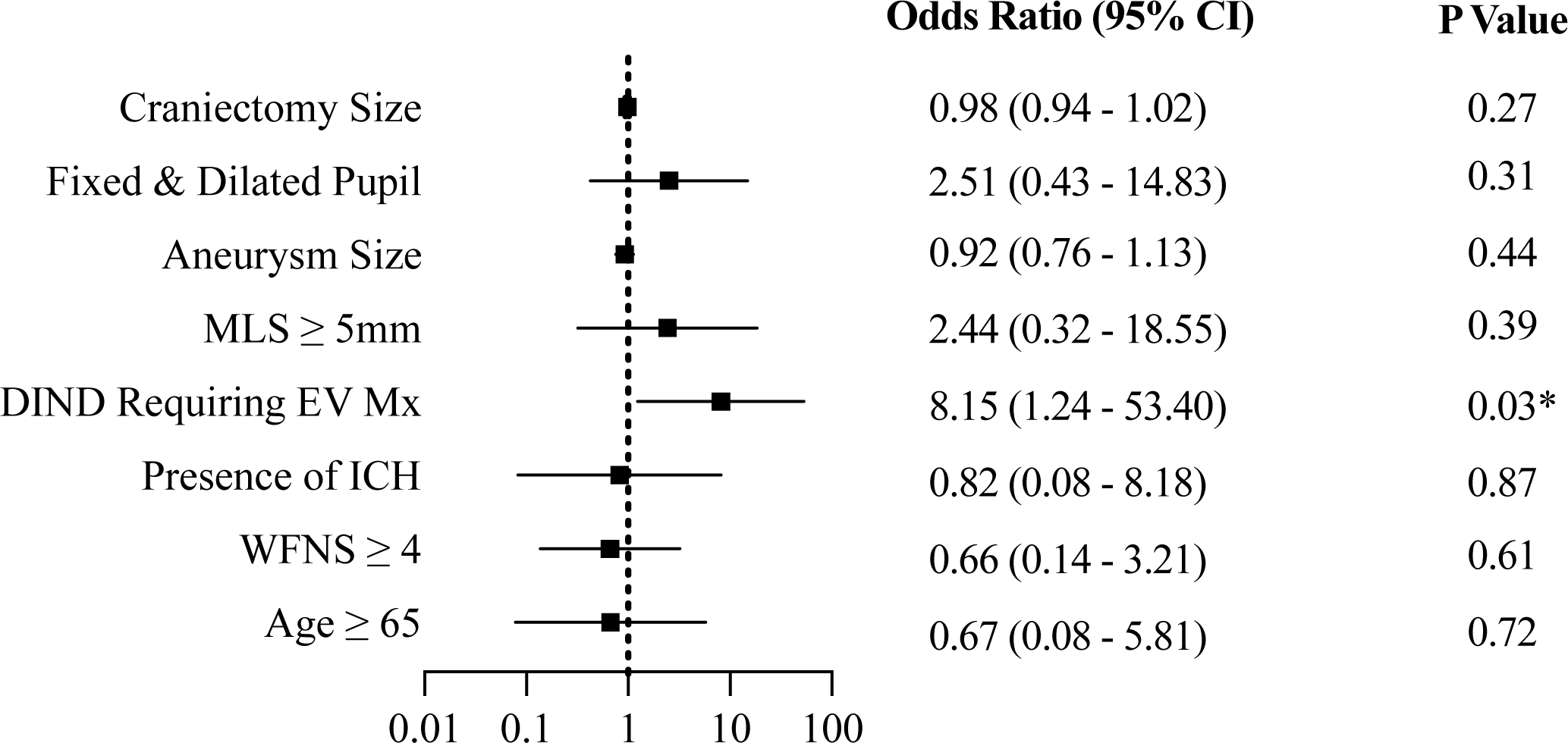
Independent Predictors of Positive NIS-SOM Score. MLS: Midline Shift; DIND: Delayed Ischaemic Neurological Deficit; EV: Endovascular; Mx: Management; ICH: Intracerebral Haemorrhage; WFNS: World Federation of Neurosurgical Societies

## Discussion

Previously published investigations of the role of DC for patients with aSAH has yielded varied experience with respect to predictors of requiring decompressive surgery and outcome. ^12,21^ This analysis of a contemporary Australian aSAH cohort undergoing DC is consistent with some, and contradicts others, of these publications.

### Overall Outcome

Unsurprisingly, and concordant with a number of previous studies, ^12,21,22^ we observed that patients undergoing DC were significantly more likely to have an unfavourable outcome than those who did not. This is likely to reflect the higher rates of poor presenting clinical and radiological grade among those undergoing decompressive surgery and has led some ^23^ to recommend caution before embarking upon this treatment. This position is supported by findings reported by Goedemans et al ^24^, who observed that 83% of 53 patients treated with DC experienced an unfavourable outcome, and Veldeman et al, ^23^ who reported that none of the 63 patients in their series were discharged with a favourable outcome, and only 15.9% of patients were independent at one year. Similarly, in their meta-analysis of 15 studies including patients with poor-grade (WFNS 4 or 5) SAH, Alotaibi et al ^12^ found an overall rate of poor outcome of 61.2% and death of 27.8% at one year. Despite this, the results of these analyses are confounded by the fact that many patients presented in poor neurological condition and underwent secondary DC for the management of intractable intracranial hypertension, and it is generally accepted that a greater proportion of well-selected patients can experience a favourable outcome after decompressive surgery. ^2,25–27^ Consistent with this, among a cohort of patients where 93.5% underwent pre-emptive DC and 41.3% had a WFNS grade <4, we found that almost two-thirds of patients achieved a favourable outcome at final follow-up. These findings are similar to the 62% of patients with an excellent or good outcome in a series published by Smith and colleagues, ^27^ and the 56% of patients with a Hunt & Kosnik grade IV aSAH who achieved a favourable outcome after DC reported by Otani et al ^2^.

### Propensity Matching

In an attempt to mitigate the selection bias inherent in previous investigations of DC among those with aSAH, we sought to compare outcomes between patients undergoing DC with a propensity-matched cohort who did not. Patients were well matched for age, gender and the proportion with a delayed ischaemic neurological deficit requiring endovascular angioplasty, but were not well matched for presenting WFNS grade. Patients undergoing decompressive craniectomy fared significantly worse than their matched counterparts at all endpoints, which appeared disproportionate to the baseline difference in WFNS grade. This finding contradicts Alotaibi and colleagues’ ^12^ meta-analysis of the three previously published studies with matched control groups, ^26,28,29^ which reported no difference in poor outcome at early follow-up (OR 1.3, 95% CI 0.8 – 5.2, p=0.2), 12-month follow-up (OR 1.7, 95% CI 0.4 - 4.3, p=0.2), early mortality (OR 0.6, 95% CI 0.3 - 1.3, p=0.2) or 12-month mortality (OR 1.1, 95% CI 0.6 – 2.1, p=0.8). However, of the included studies, D’Ambrozio et al ^28^ and Zhao et al ^26^ both included fewer than 25 patients in the DC arm, and there were statistically significant differences in the age and presenting GCS between the study and control groups in the latter investigation. Brandecker and colleagues ^25^ subsequently performed a comparison of 38 poor-grade aSAH patients treated with DC and 25 matched controls and also observed no difference in favourable outcome or mortality at 2-years. It is uncertain whether the worsened outcomes associated with DC in our cohort relates to inadequate matching or lack of benefit. Further and larger studies with propensity-matched controls may inform a judgement about the benefit, or otherwise, of DC among those with aSAH. However, ultimately, randomized controlled trials (such as the recently initiated PICASSO trial) will be required to provide more definitive evidence. ^30^

### Predictors of Undergoing DC

As is the case with malignant cerebral infarction, ^15^ there is growing recognition that earlier DC is associated with improved outcomes among selected patients with aSAH when compared with delayed decompression. ^4,12,14^ With this in mind, there has been significant interest in the identification of risk-factors associated with delayed intracranial hypertension to permit earlier selection of patients who are likely to require DC. In their recently published meta-analysis of 28 studies involving 2788 patients, Oppong and colleagues observed that younger individuals (p <0.00001; MD = −3.6 years; 95% CI −5.2 to −2.1), those with poor grade aSAH (p<0.00001; OR 4.8; 95% CI 2.9-8.0) and patients with associated ICH (p<0.00001; OR = 6.6; 95% CI 4.0 – 11.0) were more likely to receive DC. ^21^ Similarly, Veldeman et al recently reported that younger age, higher radiological grade, increasing aneurysm size and anterior circulation location were all independently associated with delayed intracranial hypertension requiring DC in multivariable logistic regression. ^23^ In our analysis, the presence of ICH at admission, and increasing pre-operative MLS, were both predictive of undergoing DC. Age, WFNS grade, vasospasm requiring endovascular management, presence of unilateral fixed and dilated pupil, aneurysm size and aneurysm location were all non-predictive. It is important to recognise the distinction between these variables representing predictors of those who underwent DC, rather than necessarily being predictors of requiring a DC. They are significantly affected by selection bias, with large discrepancies in the predictive value of particular data-points reflecting inter-institutional variability in indications for recommending DC, the proportion of patients undergoing up-front rather than delayed DC, preferences for surgical or endovascular management of the ruptured aneurysm and preferences regarding the appropriateness of aggressive surgical management in favour of palliative treatment. In an attempt to mitigate this, we sought to identify characteristics that were particular to a subgroup of those who underwent a DC at the time of their index operation, but also required post-operative neuromuscular blockade, hyperosmolar therapy or barbiturate coma for management of ongoing intracranial hypertension. Only a little over one-quarter of included patients met these criteria, which left an insufficiently powered sample to complete this analysis. However, we believe that it provides the best methodology to identify predictors of delayed intracranial hypertension among those undergoing early surgery and recommend further exploration in the future.

Recognising the potential value of early prediction of requiring DC among patients with aSAH, Jabbarli et al ^22^ recently devised the PRESSURE score, which is designed to assist in predicting patients who will require primary or secondary DC after aSAH. In their analysis, 63.7% of patients with a PRESSURE score of ≥ 6 required a decompressive craniectomy, compared with 12% of patients with a score of < 6 (OR=12.9 (95% CI 9.3-17.8), p<0.0001) and at-risk patients who underwent early DC (within 24hrs of admission) had a lower risk of unfavourable outcome [OR=0.5 (95% CI 0.2-0.9), p=0.03] and in-hospital mortality [OR=0.3 (95% CI 0.1 - 0.8), p=0.02] than those undergoing late or no DC. From this, they inferred that clinicians should consider the use of the PRESSURE score to guide prophylactic craniectomy for patients who are at risk of requiring this surgery.

While we agree that early DC is more likely to confer a benefit than delayed surgery, and that a predictive tool could have tremendous clinical utility, the PRESSURE score suffers from a number of significant shortcomings. Firstly, the cut-off score of 6 was not highly discriminatory, with over one-third of patients in the ‘high risk’ category not undergoing a DC in the studied populations. Secondly, one of the domains required assessment of the patient’s digital subtraction angiogram for early vasospasm, which is unlikely to be performed at presentation among those in extremis from an intracerebral haematoma. Given that these patients are at highest risk of requiring a DC, ^24,31^ a score that does not assist in the assessment of whether the surgeon should perform a primary DC at the time of the index surgery is of limited practical utility. With this in mind, we believe that there remains scope for further research to identify additional factors that may assist in the prediction of patients who are likely to require DC after aSAH.

### Outcome Prediction

We observed that WFNS grade ≥ 4 was associated with unfavourable outcome at follow-up, and that patients requiring endovascular vasospasm management were more likely to have an unfavourable outcome at follow-up and a positive NIS-SOM result. This is consistent with the findings from Oppong and colleagues’ meta-analysis, ^21^ which reported that younger age and less severe grade were most predictive of favourable outcome. Consistent with findings published by Otani and colleagues, ^2^ but contrary to those observed in a small series published by Kazumata et al, ^32^ we did not observe any association between extent of pre-operative MLS or haematoma volume.

As has been observed in the traumatic brain injury ^18,33,34^ and ischaemic stroke ^35^ context, increased DC size has been associated with improved functional outcome among some patients with aSAH. ^36^ However, this association has not been observed in all studies, ^4,37^ including our own, and requires further investigation.

### Limitations

It is important that our research be interpreted in the context of its limitations. First, as with all previously published research on this topic, our work is subject to selection bias, with the indication, technique and timing of DC at the discretion of the treating neurosurgeon. We have attempted to mitigate this by performing subgroup analyses on patients undergoing DC and requiring aggressive post-operative medical management of elevated ICP, as well as proceeding with comparison of those undergoing DC with a propensity-matched cohort who did not. While these analyses suffered from the shortcomings discussed above, we believe that similar future analyses will provide the best indication of those who may develop intracranial hypertension and, potentially, benefit from early decompressive surgery. Second, despite being relatively large in comparison with many previously published reports, the infrequency of intracranial hypertension following aSAH meant that our sample size was modest and limited the utility of some of the prediction models. Last, the proportion of patients undergoing secondary DC was small, limiting conclusions in this cohort.

## Conclusion

Decompressive hemicraniectomy remains a controversial intervention for the management of established or anticipated intracranial hypertension among patients with aSAH. Nevertheless, our data suggest that it can be performed with acceptable rates of morbidity and mortality, particularly among patients who present with lower grade haemorrhages. A judgement regarding the superiority, or otherwise, of DC compared with structured medical management of intracranial hypertension remains elusive and requires clarification in future propensity-matched or, ideally, randomized research. Similarly, although previous research from the aSAH and ischaemic stroke literature would suggest that earlier decompressive surgery is associated with superior outcome when compared with rescue DC, further investigation is required to identify easily measurable predictors of delayed intracranial hypertension and poor outcome in this patient cohort.

## Data Availability

The data that support the findings of this study are available on reasonable request from the corresponding author, TJO

## Acknowledgements

The authors would like to thank Dr Aye-Aye Gyi for her assistance in data extraction for this research

